# A comparison of random mixing in a structured agent-based model with empirical contact survey data

**DOI:** 10.1101/2025.09.18.25336044

**Authors:** Janik Suer, Johannes Ponge, Michael Brüggemann, Jan Pablo Burgard, Vitaly Belik, Bernd Hellingrath, Alejandra Rincón Hidalgo, Andrzej K. Jarynowski, Richard Pastor, Huynh Thi Phuong, Steven Schulz, Ashish Thampi, Chao Xu, Marlli Zambrano, Rafael Mikolajczyk, André Karch, Veronika K. Jaeger, the OptimAgent Consortium

## Abstract

Agent-based models (ABMs) are powerful tools for simulating disease spread, relying on individual-level representations and interaction rules from which emergent dynamics arise. These rules need to be accurately specified as minor differences can lead to vastly different disease dynamics. An important component in ABMs is the contact behaviour. To decrease the computational complexity the contact behaviour is often assumed as random mixing within settings. Here, individuals randomly contact other individuals associated with the same settings, such as colleagues in their workplace, where the setting associations are based on empirical data such as census data. However, the validity of the random mixing assumption within settings remains unclear. We address this gap by comparing the contact structure in a large-scale ABM (GEMS) with empirical contact survey data (COVIMOD). We compare the age contact matrices for households, schools, workplaces, all remaining contact settings combined, and all contacts combined. This includes calculating the difference matrix and the sum of squared errors (SSE), i.e. the element-wise squared difference. Our results demonstrate that random mixing in settings generated based on known age-compositions like households (SSE:0.7(95%CI0.4–0.9)), schools (SSE:0.7(95%CI:0.3–1.1)) and workplaces (SSE:0.5(95%CI:0.2-0.7)), can capture the basic interaction patterns. However, it fails to account for age-related variations in contact numbers, leading to discrepancies between the simulated and observed contact behaviour. The largest differences arise for contacts outside of households, schools and workplaces (SSE:3.8(95%CI:1.2–6.5)), due to the model’s structure. These contacts are modelled as random regional contacts not capturing the age-structured behaviour observed in COVIMOD. We conclude that random mixing in accurately defined settings provides an approximation for contact structures in settings where the age-structure of associated individuals is similar to the observed contact structure. For settings where the age-structure deviates from the contact structure, advanced methods are required to represent real-world contact structures.

**Author Summary:** Infectious disease modelling is a prominent tool for understanding disease spread and evaluating potential countermeasures. Agent-based models simulate the behaviour of individuals based on a set of simple rules and allow for a detailed representation of disease transmissions. One essential rule is the contact behaviour of individuals, which determines the potential transmission pathways of pathogens. Agent-based models often assume simple random-mixing of individuals in the locations they are associated with, such as their households or workplaces. We investigate if this assumption is appropriate by comparing the contact structure simulated in the agent-based model GEMS with the contact data gathered through the Germany-wide COVIMOD survey. We find that the random mixing can serve as an approximation for settings such as household, schools and workplaces where the contact structure is similar to the age structure of the individuals present in the setting. However, discrepancies arise as random mixing cannot account for the differing number of contacts by age or household size. We identified the most discrepancies for contacts outside of the household, school and workplace. Here, the highly age-structured contact behaviour observed in COVIMOD strongly deviates from the random mixing of all ages observed in GEMS.

## Introduction

Understanding the complex dynamics of disease spread requires tools capable of capturing individual-level behaviours and interactions. Agent-based models (ABMs) provide this capability, enabling the simulation of epidemics in realistic populations. The accuracy of ABMs for modelling disease spread relies on the accurate specification of individual behaviours, in particular the contact behaviour. Depending on the type of pathogen considered, the contacts that need to be considered in the simulation can vary. For instance, when modelling respiratory infections such as COVID-19, face-to-face contacts between individuals arising from, e.g., conversing with or sitting near other people are the most important types of contact that need to be considered.

The contact behaviour of individuals has been studied frequently on different scales in various populations [1–4]. Popular studies on individuals contact behaviour include the POLYMOD survey [2], which assessed the contact behaviour of individuals in multiple European countries and has been used to parametrise different infectious disease models [5–8]. However, as the contact behaviour of individuals has changed over the years, due to technological and societal changes, newer studies should be considered [9]. The COVIMOD study is one such recent study that assessed the contact behaviour of individuals during and shortly after the COVID-19 pandemic in Germany [4,10–12].

While ABMs can theoretically model contact behaviour with arbitrary complexity, computational limitations require the use of simplified approaches, to enable efficient simulation. These considerations are particularly relevant for models that aim to simulate large populations [6,13–15]. One of such models is GEMS (German Epidemic Microsimulation System), which facilitates the simulation of disease dynamics in the entire population of Germany [14,15]. GEMS assumes random mixing of the individuals associated with the same setting to model their contacts. These settings correspond to the individuals’ household, workplace, school and municipality. Therefore, the simulated contact structure does not correspond to random mixing of the entire population but to a random mixing of the individuals associated with the same setting, imposing the age-structure of the setting as its contact structure. The age-structure of the settings is created during the development of the synthetic population. Here, demographic and setting specific data such as census data and workplace data are being used to create individuals and associate them with the appropriate settings. Using a random mixing in the settings yields a computationally efficient model but may not be capable of representing the nuanced contact structure observed in the real world. This raises the central question: *Can the random mixing assumption within settings reproduce the contact behaviour observed in empirical surveys?*

To address this question, we compare the contact structure generated by GEMS with the empirical data from the COVIMOD survey. The comparison involves three steps. First, we extract the contact matrices from GEMS and the COVIMOD survey. Second, we use the setting specific contact matrices derived from the COVIMOD data to calibrate the contact behaviour in GEMS, by minimizing the difference between the simulated and measured contact matrices.

Third, we investigate the remaining structural differences and identify their origins. This analysis is performed for the household, workplace, school and all contacts outside of the households, workplaces and schools, henceforth denoted as other contacts as well as all contacts combined. Our findings suggest that while random mixing in GEMS leads to contact structures that, in general, align well with COVIMOD data, some structural differences remain, highlighting the potential benefits of implementing a more refined contact behaviour in ABMs.

## Methods

### Data

The COVIMOD study is a longitudinal cohort study designed to capture the contact behaviour of the German population during and after the COVID-19 pandemic. The study consists of 36 waves during which a demographically representative sample of the German population completed a questionnaire regarding themselves and their contacts on the preceding day. To capture child contacts, a subset of adults with children under the age of 18 completed the questionnaire as proxies for their children. Further details can be found elsewhere [4,10–12]. Since we want to consider the contact behaviour under non-restrictive conditions, we use the final three survey waves (wave 34 – 36), conducted between November 2022 and March 2023. These waves capture the contacts of 7,481 participants, who reported a total of 23,146 contacts categorized as household (8,056), workplace (4,495), school (2,370) and other contacts (8,224), i.e., contact the participant had in their household, workplace, school or in any other place, respectively. Note that while COVIMOD includes more contact settings, we group them together as “other contacts” to allow for a comparison with GEMS. Furthermore, within COVIMOD the same participants were asked for their contacts in multiple waves. We treat these multiple responses as independent participants.

Prior to the analysis, missing participants’ age data was imputed. For participants that only supplied their age-group the exact age was imputed using a weighted sampling based on the 2022 age-distribution in Germany [16]. Using the same procedure, we imputed the ages of group contacts, i.e., contacts where participants reported the number of individuals participating in the group contact and their collective age group.

Contact matrices for the four contact categories (household, workplace, school and other) were generated from the COVIMOD data using the socialmixr package [17], a package for the R programming language R [18] commonly used for creating contact matrices based on contact survey data. The entries m_ij_ of these contact matrices represent the average number of contacts reported by individuals in age group A_i_ with individuals in age group A_j_ during a 24-hour period in the given setting. Age groups were defined using five-year right-open intervals. To account for variations in contact behaviour between weekends and weekdays, contacts were weighted based on whether they were reported on a weekday (weight prop. to 5/7) or on a weekend (weight prop. to 2/7) to obtain the contact behaviour for an “average” day.

Contacts between people are in general symmetric, meaning if person A has a contact with person B, B also has a contact with A. This symmetry can be expressed as:

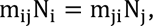

where m_ij_ and m_ji_represent the corresponding entries of the contact matrix and N_i_ and N_j_ the size of the age-groups i and j, respectively. This symmetry was used in the socialmixr package when constructing the contact matrices for household contacts, other contacts and all combined contacts. However, this condition does not hold for schools and workplaces due to reporting differences. For example, a teacher-student contact will be reported as a workplace contact by the teacher and a school contact by the student. Consequently, the symmetry condition was not employed for the contact matrices of workplaces and schools. Even for household and other contacts, the condition only holds approximately as certain interactions, such as contacts of the elderly with visiting nurses at home, will be asymmetrical, i.e., reported as contacts at home and work, respectively. For household contacts, the symmetry was employed to mitigate the underrepresentation of adults living with children in the survey, as participants with children were asked to fill out the form as proxies for their children and not for themselves therefore disturbing the originally representative sampling.

### GEMS

GEMS (German Epidemic Microsimulation System) is an agent-based model that allows the simulation of disease spread within the entire population of Germany. In GEMS individuals are associated with settings where they can have contacts [14]. Therefore, the potential contact partners of a person are the other members of the settings the person is associated with. The settings included in GEMS are households, workplaces, schools and municipalities, with the latter being used to simulate all contacts outside of households, workplaces and schools. Schools and workplaces are implemented using four hierarchical substructures. For the schools, these substructures are the school complex, schools, school years and classes, where each includes at least one of the latter. Similarly, the workplaces are implemented as workplace sites, workplaces, departments and offices. In each of these settings, including the substructures, individuals are assumed to be mixing randomly such that each contact is equally likely.

The contact sampling for a specific individual I in the setting S_T_ of type T works in two steps. First, the number of contacts N_C_the individual I will have is drawn from a Poisson distribution with mean λ_T_, depending on the type T of the setting S_T_. Second, from the individuals present in the setting S_T_ excluding individual I, N_C_ individuals are randomly drawn with replacement and recorded as contacts. For our analysis contacts are drawn for randomly selected individuals in the population until the total number of 10^7^ recorded contacts are reached. This large number of contacts was chosen as a cut-off to assure that uncertainties of the simulated contact behaviour are negligible for further analyses. For every selected individual all contacts they have during one day are sampled. Based on the age of the selected and sampled individuals the simulated contact matrix can be constructed.

The structure of contacts in GEMS is a result of the association of individuals to settings. These associations are determined during the generation of the Gesyland population [19], a synthetic population that represents various characteristics of the German population. The locations and types of settings, such as households, schools or workplaces, are obtained from OpenStreetMap [20]. They are provided with characteristics derived from the 2011 German census on a 100m square grid [21], including the household size distribution. Individuals are created based on demographic, education, occupation and health related variables obtained from the German microcensus [22] and the German health survey GEDA [23]. They are assigned to households, schools and workplaces based on individual characteristics and the interregional commuting structure derived from commuting tables [24].

While we use GEMS for our comparison, other ABMs follow the same methodology. Using an accurate description for the population and its association to specific location which allow for random contacts of its members [6,25–27]. Models that employ this structure allow for a simplified computation of contacts by shifting the work from the contact sampling during the simulation to the creation of an accurate synthetic population before the simulation.

### Comparison

As the initial model parametrisation, the average number of contacts per day is set to one, i.e., λ_T_ = 1 for all setting types T in the simulation. Based on the simulated contact matrix 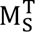 and the contact matrix derived from COVIMOD data 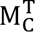 the average number of contacts λ_T_can be calibrated as follows. Note that to simplify the notation, from now on we drop the setting type index T, while still referring to setting type specific variables. Since the entries in the contact matrix scale linearly with the defined average number of contacts λ, scaling the average number of contacts by α, such that λ′ = αλ results in a linearly scaled contact matrix 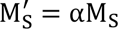. To describe the difference between the two contact matrices, we use the sum of squared errors (SSE), placing more emphasis on large deviations than small ones. The SSE for the survey-based contact matrix M_C_ and the linearly scaled simulated contact matrix 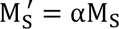 can be expressed as:

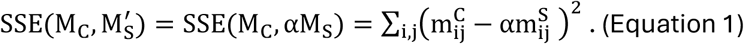

Although the unit of the SSE is number of contacts squared, we will treat it as a dimensionless number for brevity. Equation 1 is minimized at:

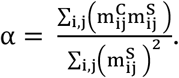

Calculating α yields the optimal average number of contacts for settings such as households, without further substructure. However, for settings which have substructures, i.e., schools and workplaces, the simulated contact matrix is equal to the sum of the contact matrices of the substructures which can be scaled individually. Therefore, the SSE can be expressed as:

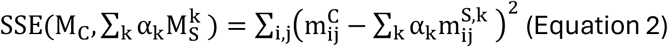

Here, 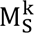 corresponds to the contact matrix of the k-th substructure, with their respective scaling factors α_k_. In the following the α_k_ values for the substructures are obtained by numerically minimizing the Equation 2 using Nelder-Mead algorithm implemented by the Optim package [28] in Julia [29].

We compare the contact matrices for households, schools, workplaces, “other contact” and all contacts combined simulated in GEMS and based on the COVIMOD survey data visually and quantitatively. For visual comparison, the differences between the simulated and survey-based contact matrices are displayed. Here, we show the differences in matrix form to be able to identify structural under- and overestimation of contacts between specific age-groups. For quantitative comparison, the minimal SSE is used as a measure of difference between two contact matrices. We calculate the 95% confidence intervals (CIs) of the minimal SSE by bootstrapping with respect to the participants of the COVIMOD survey. The uncertainties of contact matrices estimated for GEMS are negligible due to the large sample size, which was ascertained by also bootstrapping with respect to the simulated individuals in GEMS, yielding CIs identical to the ones calculated by just bootstrapping the COVIMOD participants.

## Results

This section presents the comparison of the contact matrices derived from the COVIMOD survey and the GEMS model separated into the contacts in households, schools, workplaces and other contacts. Lastly the combined contact structure will be compared.

### Contacts in the setting “Household”

Figure 1 (A) displays the contact matrix for the household setting from the COVIMOD survey. The matrix shows a prominent diagonal line, corresponding to age-assortative mixing due to individuals of the same age living together as couples, siblings, or roommates. Additionally, two off-diagonal lines are present. These lines represent intergenerational contacts of families living together, such as the household contacts of children with their parents.

**Figure 1:**
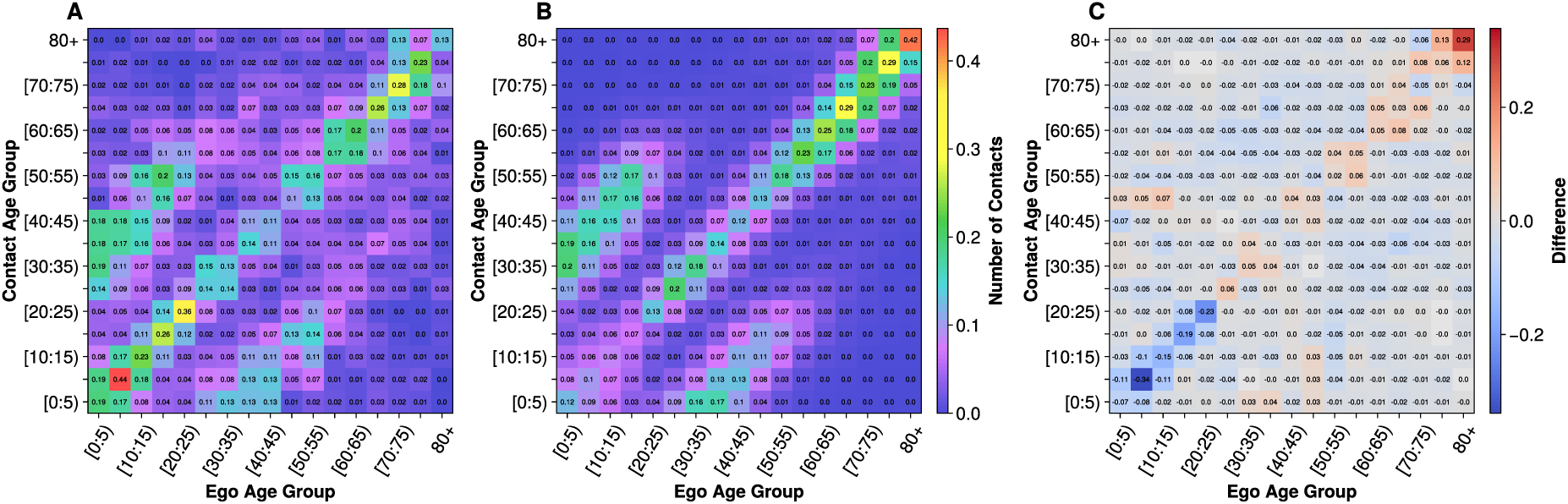
Comparison of the age x age contact matrices for household contacts. (A) Contact matrix for the household contacts derived from COVIMOD data. (B) Calibrated household contact matrix simulated in GEMS. (C) Difference between the GEMS and COVIMOD contact matrices (B – A) negative values correspond to an underrepresentation of contacts in GEMS compared to COVIMOD.

The calibrated household contact matrix generated in GEMS is shown in Figure 1 (B). It displays a similar structure as the COVIMOD data, with a main diagonal representing age assortative living conditions and off-diagonal lines representing inter-generational contacts. In GEMS, the oldest age group (80+) shows the highest number of contacts within their own group, whereas in COVIMOD the children aged 5-10 exhibit the highest number of contacts. The difference is a result of the implemented form of random mixing, which assumes the same number of daily contacts for all individuals. This causes individuals living in larger households with different age-groups, such as families, to have a contact pattern distributed over the present age-groups. At the same time, individuals living in small households with few or only one other individual have the same number of contacts as individuals living in families but with fewer different age groups.

Calibrating the contact sampling in GEMS with the COVIMOD contact matrix, by minimizing SSEs between the contact matrices, results in an average number of household contacts of 𝜆_𝐻_ = 0.92. The difference between the calibrated GEMS contact matrix and the COVIMOD contact matrix is displayed in Figure 1 (C). The biggest differences can be seen in for children and young adults under the age of 25. Here, the number of contacts people have with their own age group and neighbouring age groups is underestimated, most noticeably for the [5,10) age group. At the same time the age assortative contacts of the 80+ age group are overrepresented. These differences highlight the limitation of the random mixing implemented in GEMS, which assumes the same number of contacts for all individuals. While the model captures the overall structure of household contacts it fails to account for the high number of contacts among children and young adults, as well as the low social activity of the elderly. Addressing these issues requires household-size-specific or age-specific contact behaviour to be implemented, since simply adjusting the average number of contacts would result in a linear scaling for all age-groups.

### Contacts in the Setting “School”

Figure 2 (A) shows the contact matrix for school contacts in the COVIMOD survey. Again, the age-assortative mixing is visible as a diagonal line. This mixing is facilitated by the structure of the school system, where children are in school classes with other children of similar age. The contact matrix also displays minor off-diagonal elements, which might stem from the choice of age group size for the youngest age groups. In Germany, children are generally admitted to primary school at the age of six, therefore contacts between the two youngest age groups may arise from kindergarten contacts between children under five years and children over five who have not yet been admitted to school. For the [20, 25) group the off-diagonal elements likely reflect contacts within universities, which exhibit a less age-dependent structure. Beyond the age of 25 the entries in the contact matrix vanish as people are likely to have left the educational system. Additionally, a few contacts between young and middle-aged individuals can be observed reflecting contacts of students with teachers.

**Figure 2:**
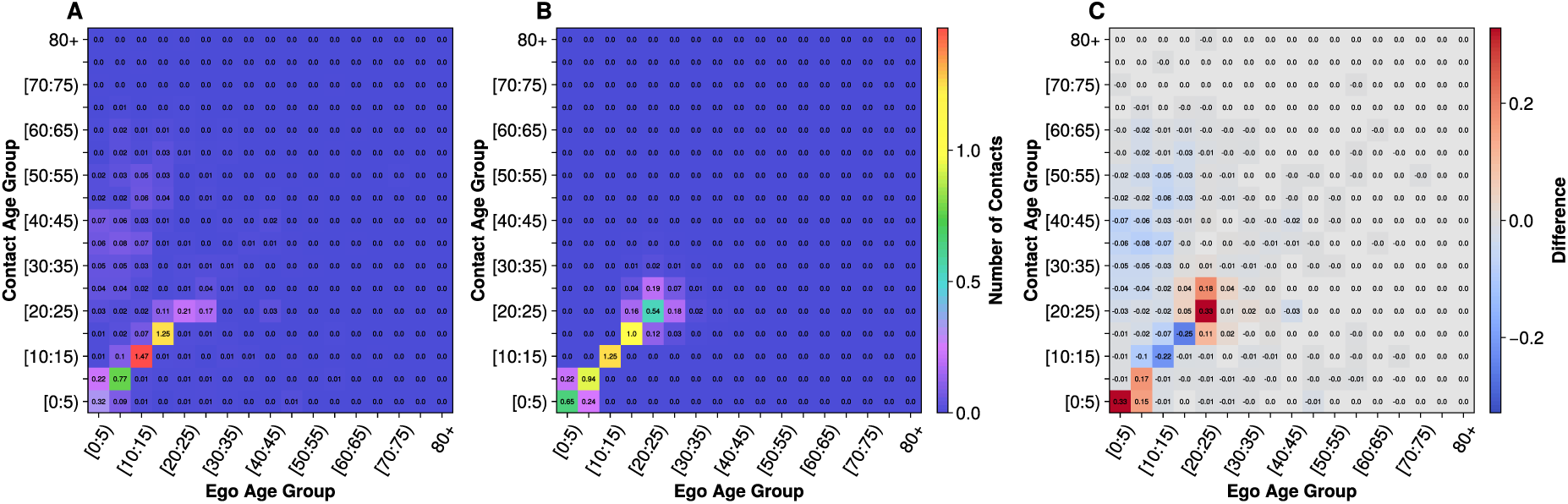
Comparison of the age x age contact matrices for school contacts. (A) Contact matrix derived from the COVIMOD survey data. (B) School contact matrix derived from the simulated contact behaviour in GEMS. (C) Difference between the GEMS and COVIMOD contact matrices (B – A) negative values correspond to an underrepresentation of contacts in GEMS compared to COVIMOD.

The contact matrices for the uncalibrated substructures of the school in GEMS are displayed in Figure 6 in the Appendix. Here, the school classes and school years represent highly age-assortative mixing while schools and school complexes also capture contacts between age-groups. Calibration of the average number of contacts for the substructures yields: 𝜆*_schoolyear_*= 1.26 and 𝜆*_class_* = 𝜆*_school_* = *𝜆_schoolcomplex_* ≈ 0. Thus, the age structure of school years alone most closely replicates the contact structure observed in COVIMOD. The calibrated GEMS school contact matrix is displayed in Figure 2 (B).

The difference between the calibrated GEMS and COVIMOD school contact matrices is displayed in Figure 2 (C). In GEMS, the age assortative contacts of the age group [10,15) are significantly underestimated, while those of the youngest age group are overestimated. Besides the age-assortative contacts the mixing between neighbouring age-groups is overestimated for the young and older age-groups while it is underestimated for the groups in between, highlighting the limitation of the employed form random sampling. As with the household contacts mitigating the under- and overestimation requires an age-dependent contact sampling to describe differences in social activity. Note that a class-dependent social activity could also be defined as the class is directly related to the age. Additionally, the teacher-pupil contacts are not captured in GEMS due to the employed setting structure. This does not include teachers in the school setting but rather treats them as part of the regular workforce and assigns them a workplace.

### Contacts in the Setting “Workplace”

Figure 3 (A) displays the contact matrix for workplace contacts obtained from the COVIMOD survey. Workplace contacts emerge in the age group [15,20) and continue until the age group of [65,70) with only some contacts for older age groups, reflecting the typical retirement age of 65 years. Within this working population the number of contacts strongly fluctuates. Additionally, contacts of working individuals with elderly people and children can be observed, likely representing care providers and teachers, respectively.

**Figure 3:**
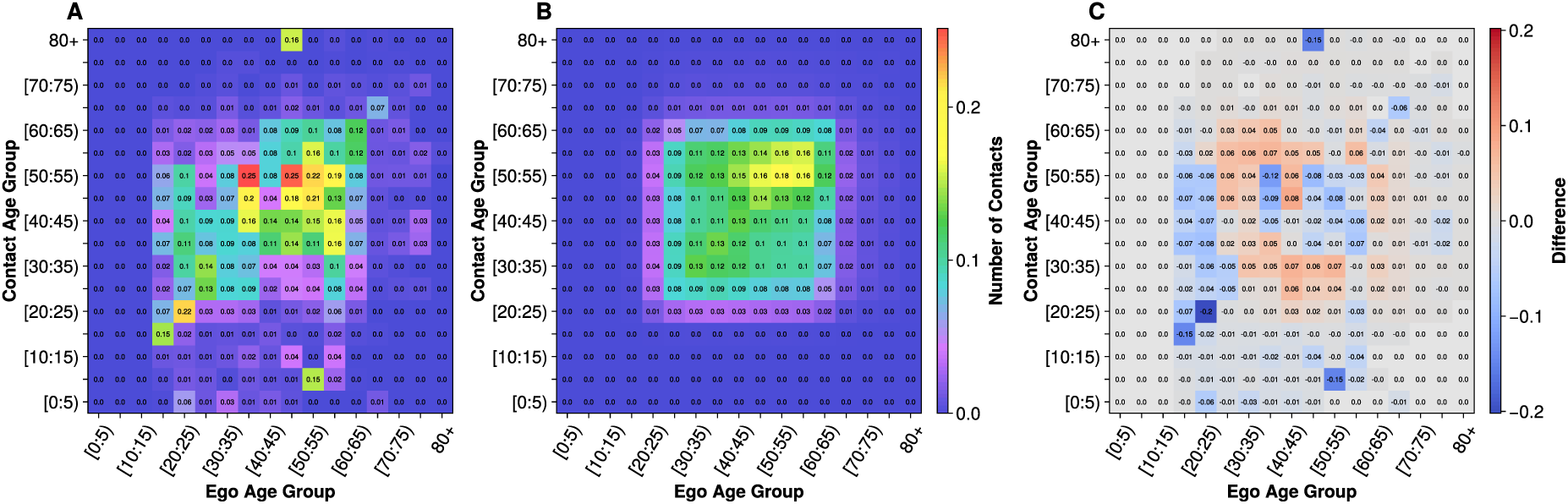
Comparison of the age x age contact matrices for workplace contacts. (A) Contact matrix for the COVIMOD workplace contacts including asymmetric contacts, e.g., teacher-pupil contacts. (B) Workplace contact matrix based on the contact behaviour simulated in GEMS. (C) Difference between the GEMS and COVIMOD workplace contact matrices (B – A) negative values correspond to an underrepresentation of contacts in GEMS compared to COVIMOD.

The uncalibrated contact matrices for the four workplace substructures are presented in Figure 7 in the Appendix. As all substructures have a similar age structure, i.e., the age-constellation of workers in an office are on average not different from the age-constellation of entire companies, all substructures also exhibit a similar contact structure. The calibration of the contact numbers of the substructures with the COVIMOD data results in 𝜆*_workplace_* = 0.29, 𝜆*_department_* = 0.81 and 𝜆*_office_* = 𝜆*_workplacesite_* ≈ 0. The calibrated GEMS workplace contact matrix is displayed in Figure 3 (B). This matrix consists of a rectangular structure encompassing the working-age population starting from the age group [20,25) and ending with the [60,65) group, with a low number of contacts for the [65,70) group. The internal structure of the rectangle reflects the age distribution of the working population with most contacts of all working ages occurring with the [50,55) group, since it corresponds to the largest group in the working population.

The difference between the working contacts in COVIMOD and GEMS is shown in Figure 3 (C). It varies between over- and underestimation, due to the large variations in the COVIMOD data that can be observed in Figure 3 (A). A systematic underestimation of work contacts occurs for the [15,20) group, which only exhibits very few contacts in GEMS while COVIMOD shows a similar contribution as for older age-groups. Additionally, asymmetric contacts, i.e., contacts of the working population with individuals outside of this group, are underrepresented since GEMS is unable to represent them.

### Contacts in Other Settings

Figure 4 (A) displays the COVIMOD contact matrix for the other contacts. The highest number of contacts occur along the diagonal for young adults, highlighting their age-assortative mixing. Additionally, off-diagonal elements are present, reflecting the interactions between parents and children. The diagonal line exists for all age groups highlighting the age-assortative mixing of all age-groups. However, especially the middle-aged individuals show a higher contact diversity, interacting with a wider range of age groups. Contact structure becomes more age-assortative again with increasing age.

**Figure 4:**
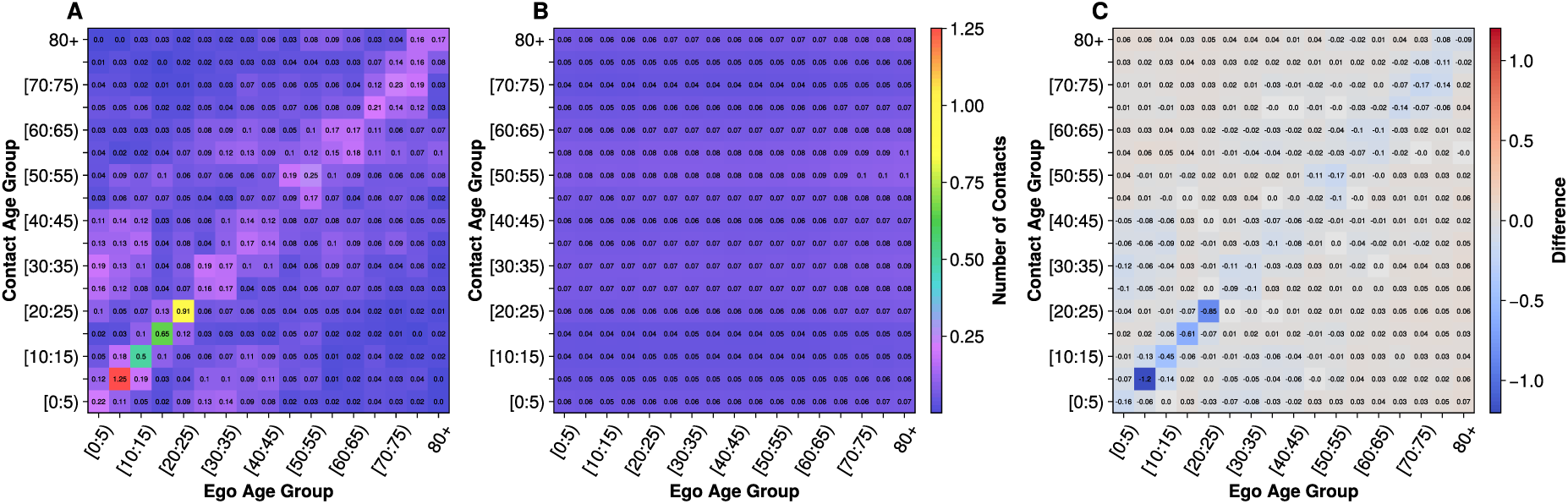
Comparison of the age x age contact matrices for the other contacts. (A) Contact matrix for the other contacts derived from the COVIMOD contact survey. (B) Contact matrix for other contacts based on the contact behaviour simulated in GEMS. (C) Difference between the GEMS and COVIMOD other contact matrices (B – A) negative values correspond to an underrepresentation of contacts in GEMS compared to COVIMOD.

Figure 4 (B) displays the contact matrix obtained for the other contacts in GEMS. In GEMS, these contacts correspond to random mixing within the municipalities, resulting in the likelihood of a contact with an age group to be equal to the share of individuals in the municipality in this age-group. Consequently, the observed contact structure in GEMS corresponds to the overall age structure in the municipalities, such that a cross section for any age group would yield the age distribution of the population. This can be observed as horizontal lines in Figure 4 (B). Therefore, GEMS fails to capture the age assortativity and heterogeneous social activity that the COVIMOD data displays. Calibrating the other contacts in GEMS to the COVIMOD data yields 𝜆_𝑂_ = 1.19.

The difference between the calibrated contact matrix in GEMS and the COVIMOD matrix is shown in Figure 4 (C). The structure of the COVIMOD contact matrix is clearly visible in the difference matrix, highlighting the systematic underestimation of the COVIMOD contact structure by GEMS. Consequently, contacts outside of these age-groups are slightly but systematically overestimated. This highlights the inherent structural limitation using random regional contacts to represent the age-structured contacts observed in the COVIMOD data, as it is used in GEMS.

### All Contacts

Figures 5 (A) and (B) show the contact matrices for all contacts observed in COVIMOD and simulated in GEMS, respectively. The difference between these matrices is shown in Figure 5 (C). While GEMS captures the overall contact structure, it severely underestimates the age-assortative contacts of children and young adults as well as contacts between children and their parents. This difference is primarily due to the limitations of GEMS in modelling the “other contacts” as mentioned before.

**Figure 5:**
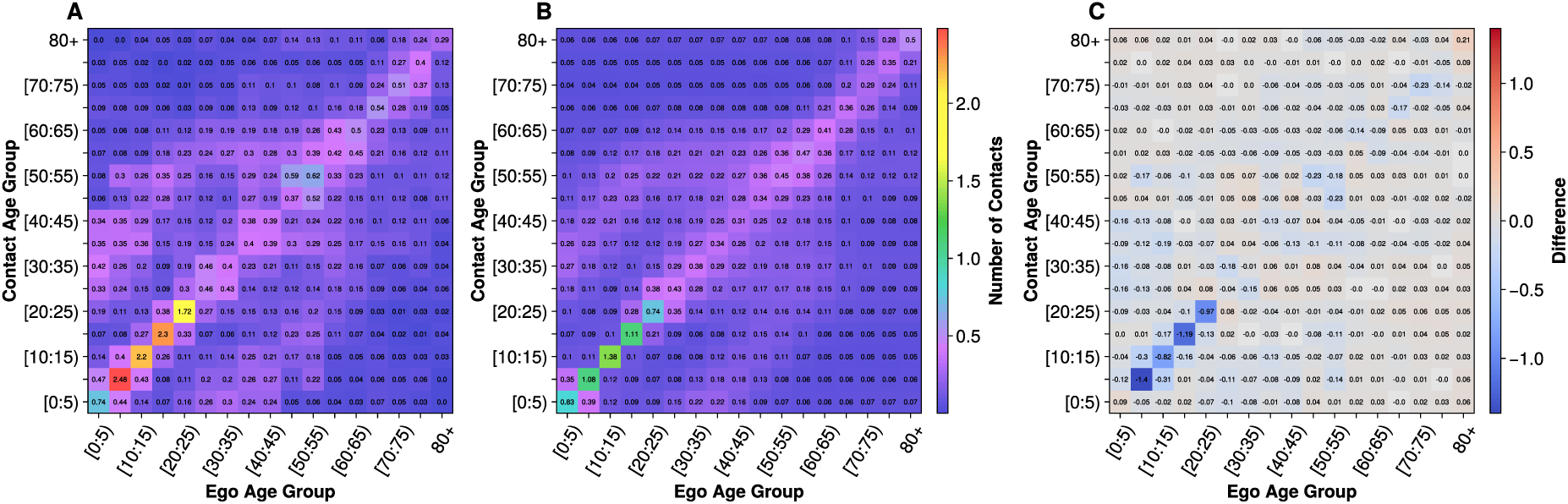
Comparison of the age x age contact matrices for all contacts combined. (A) Contact matrix for all contacts reported in the COVIMOD contact survey. (B) Combined contact matrix of all contacts simulated in GEMS. (C) Difference between the GEMS and COVIMOD contact matrices (B – A) for all combined contacts. Negative values correspond to an underrepresentation of contacts in GEMS compared to COVIMOD.

Table 1 presents the sum of squared errors (SSE) between the GEMS-generated and COVIMOD contact matrices for each contact setting calculated using Equation 1, along with the 95% confidence intervals for the obtained through bootstrapping with respect to the COVIMOD participants. Household, school and workplace contacts all show similar SSE values: 0.7 (95% CI: 0.4 to 0.9), 0.7 (0.3 to 1.1) and 0.5 (0.2 to 0.7), respectively. In contrast, the “other contacts” show a considerably higher SSE of 3.8 (1.2 to 6.5). Lastly, the deviations for all contacts lead to an SSE of 7.6 (95% CI: 2.3 to 13.0).

**Table 1:** Sum of squared errors (SSE) between contact matrices in COVIMOD and the fitted GEMS calculated using Equation 1, with 95% confidence intervals determined by bootstrapping with respect to the COVIMOD participants.

| Contact Setting | SSE (95% CI) |
| --- | --- |
| Household | 0.7 (0.4 - 0.9) |
| School | 0.7 (0.3 – 1.1) |
| Workplace | 0.5 (0.2 – 0.7) |
| Other Contacts | 3.8 (1.2 – 6.5) |
| All Contacts | 7.6 (2.3 – 13.0) |

## Discussion

In this study, we assessed if random mixing in settings could accurately represent real-world contact behaviour, by comparing the contact matrices derived from a large-scale ABM (GEMS) with empirical contact survey data (COVIMOD). Our results demonstrate that the implemented random mixing captures the basic structure of empirically assessed contact behaviour in settings with a restricted age-structure such as schools and workplaces. In these settings it is struggling to represent the age-dependent variations in social activity. Specifically, random sampling can represent the age assortative contacts in schools and households, the children-parent interactions in households, as well as the homogeneous mixing of the working population for workplace contacts. Even still, all these contacts show small deviation from the contacts observed in COVIMOD that arise from two sources. First, the implemented random mixing assumes the same average number of daily contacts for all individuals, which fails to account for heterogeneous social activity known to be age-dependent [30], thus causing mismatches especially for the children and elderly. Second, the inherent assumption of GEMS that the contacts between individuals occur only if both contact partners perform the same role, such as school contacts only including contacts between students. Therefore, contacts that occur between individuals performing different roles, e.g., teacher-student contacts, are not included. This separation of contacts by roles is not an assumption unique to GEMS, but commonly used to simplify the contact behaviour in ABMs [25–27,31]. However, the most severe mismatch arises for the other contacts. This is a result of the structure of GEMS, as the “other contacts” are modelled as random contacts in the municipality, which fails to account for the heterogeneous contact behaviour observed in COVIMOD.

These discrepancies between the contact structures implied by random mixing and COVIMOD could be reduced by employing more refined contact sampling methods. Potential approaches include a setting-size dependent number of contacts. For households, the relation of increasing number of contacts for increasing household size has been demonstrated in [32]. However, it is not obvious to which extend the underestimation of child-child contacts is a result of the lack of household size dependence or actually a result of a higher number of contacts of children with the siblings. The latter has also been shown to be a determining factor for household contacts [33]. Potentially both processes are contributing and should be represented in the model, which requires methods that include a household size dependent number of contacts and a contact probability dependent on the attributes of the potential contacts, such as their age. Similar attribute-based approaches could address the limitations in representing the other contacts. Alternatively, assuming a strong overlap between the other contacts and those occurring in schools, workplaces or households, a potential approach involves sampling additional contacts within these existing settings. Using this approach for the structure observed for the “other contacts” together with the currently used random municipality-based contacts could represent both random and structured other contacts.

This study’s limitations arise from both the COVIMOD survey and GEMS. While contact surveys such as COVIMOD are the main workhorse for studying individuals’ contact behaviour in the context of infectious diseases, they exhibit inherent limitations [34]. In particular, the recall bias likely leads to the underreporting of random contacts in COVIMOD, i.e., contacts with individuals that are not known to the participant such as contacts in public transport [2,35]. As the other contacts in GEMS are currently implemented as random mixing in the municipality, these other contacts correspond to the random contacts underreported in COVIMOD, explaining some of the discrepancies in contact behaviour. The parametrisation of the other contacts could be refined by integrating data sources better suited to capturing these random contacts, such as sensor data [34]. Furthermore, in COVIMOD, parents reported on the contacts of their children as proxies, which limits the validity of their reported contact behaviour, especially for school contacts [36]. In addition, since some of the parents were asked to fill out the questionnaire on behalf of their children and not for themselves, thus parents are slightly underrepresented in the survey data as respondents for the adult group. For the household and other contacts, the symmetry condition was applied to mitigate this shortcoming.

As an abstraction of reality, GEMS introduces further limitations arising from the Gesyland population data and the models’ structure. As the contact structures that emerge based on random mixing in settings are mainly dependent on the correct association of individuals to settings, the Gesyland population itself constitutes the main limitation. The assessment of the random mixing within settings relied on accurately defined settings that capture the real (age) constellation of settings such as household compositions. While we assume the accurate association of individuals to settings the differences of the simulated and empirical contact structures could also be partly caused by inaccurate association of settings within Gesyland. Gesyland contains various approximations of the German population, including identical occupations within workplaces. For example, the researchers only work with other researchers, not secretaries. Due to limited data, characteristics that occur in less than 1% of the population are omitted, and care units (e.g. nursing homes) and institutional units (e.g. prisons) are generated based on educated guesses. Both of these are treated as households in GEMS. Additionally, GEMS’ structure includes a static assignment based on the roles they perform, e.g. students are assigned to school. Therefore, only individuals in the same “role” can be in contact. As a result, GEMS is unable to represent asymmetric contacts such as student-teacher contacts regardless of the used contact sampling method. However, as previously mentioned, the ABMs commonly only include the symmetric contacts, making GEMS a suitable model to evaluate the random mixing assumption.

Future research should refine contact sampling methods to better represent the contact behaviour observed in the real-world, especially for studies focusing on disease dynamics in specific subgroups. It could also be helpful to use other types of contact data such as sensor data for calibration, as here random contacts are more likely to be captured in the data. Furthermore, besides the average number of contacts, which was the focus of this study, the distribution of the number of contacts (i.e. inter-individual heterogeneity) is crucial for the spread of infectious diseases and should be studied using data such as the COVIMOD survey [37]. In addition, the intra-individual heterogeneity, i.e., the daily variability in an individual’s contacts is another relevant factor for disease spread [38–40]. This heterogeneity could be represented in GEMS by using the existing substructure levels. Sampling the contacts in large substructures such as the school complex rather than the school class increases the heterogeneity of contacts as contact partners are likely to vary daily. Combining contacts in different substructures allows modelling different levels of intra-individual heterogeneity.

While these refinements may allow for a more detailed representation of contact behaviour in ABMs, their impact on the simulated disease spread remains an open question. Model development should focus on methods that are as simple as possible and as complex as necessary to represent the features relevant to the research question. Studies that investigate the case numbers on the population likely will need a less detailed contact behaviour, while studies that look at specific subgroups such as teacher or nurses will require a more detailed representation of their contact behaviour. Since contact behaviour within an ABM is often an emergent property of its structure, as shown here for the static setting association in GEMS, modelers should continuously review emerging contact behaviour. A comparison of the emergent contact behaviour to real-world data, following the methodology presented in this study, should be integrated into the development process of any ABM to identify shortcomings. This could ultimately lead to the development of more accurate ABMs.

## Data Availability

The code and data to reproduce the presented results are available at:
https://github.com/JanikSuer/contact_structure_comparison. The agent-based model
GEMS is available at: https://immidd.github.io/GEMS/.

## Acknowledgements

The OptimAgent Consortium includes: Tyll Krüger (Department for Informatics and Telecommunication, Wroclaw University of Science and Technology, Wroclaw, Poland); João Vitor Pamplona, Ralf Münnich, Soheil Shams (Department of Economic and Social Statistics, Trier University, Trier, Germany); Berit Lange, Isti Rodiah (Department of Epidemiology, Helmholtz Centre for Infection Research, Brunswick, Germany); Maren Steinmann, Sebastian Gruhn, Wolfgang Greiner (Department of Health Economics and Health Care Management, School of Public Health, Bielefeld University, Bielefeld, Germany); Wolfgang Bock (Department of Mathematics, Linnaeus University, Växjö, Sweden); Lukas Bayer (Department of Mathematics, University Koblenz, Koblenz, Germany); Sudarshan Tiwari (Department of Mathematics, RPTU Kaiserslautern, Kaiserslautern, Germany); Hannah Derwanz (Faculty of Medicine, Martin Luther University of Halle Wittenberg, Halle (Saale), Germany); Aleksandr Bryzgalov, Beryl Musundi. Johannes Horn, Julian Patzner, Kahkashan Mahreen, Myka Sarajan, Moritz Kersting (Institute for Medical Epidemiology, Biometrics and Informatics (IMEBI), Interdisciplinary Center for Health Sciences, Medical School of the Martin-Luther University Halle-Wittenberg, Halle (Saale), Germany.); Markus Scholz (Institute for Medical Informatics, Statistics and Epidemiology, University of Leipzig, Leipzig, Germany); Alexander Kuhlmann (Institute for Social Medicine and Epidemiology, University of Lübeck); André Calero Valdez, Lilian Kojan (Institute of Multimedia and Interactive Systems, University of Lübeck, Lübeck, Germany); Beate Jahn, Uwe Siebert (Institute of Public Health, Medical Decision Making and Health Technology Assessment, Department of Public Health, Health Services Research and Health Technology Assessment, UMIT - University for Health Sciences, Medical Informatics and Technology, Hall i.T., Austria); Mirjam E. Kretzschmar (Julius Center for Health Sciences and Primary Care, University Medical Center Utrecht, Utrecht, Netherlands.)

## Author Contribution

Conceptualization: Janik Suer, Veronika Jaeger, André Karch, Johannes Ponge, Rafael Mikolajczyk, Bernd Hellingrath.

Data curation: Janik Suer, Veronika Jaeger, Jan Pablo Burgard.

Formal analysis: Janik Suer, Michael Brüggemann.

Funding acquisition: André Karch, Rafael Mikolajczyk, Bernd Hellingrath, Veronika Jaeger.

Investigation: Janik Suer, Michael Brüggemann, Veronika Jaeger.

Methodology: Janik Suer, Michael Brüggemann, Johannes Ponge, Jan Pablo Burgard, Veronika Jaeger, André Karch.

Project Administration: Veronika Jaeger

Resources: André Karch, Bernd Hellingrath.

Software: Janik Suer, Michael Brüggemann, Johannes Ponge, Jan Pablo Burgard.

Supervision: Veronika Jaeger, André Karch.

Validation: Janik Suer, Michael Brüggemann, Johannes Ponge.

Visualization: Janik Suer, Michael Brüggemann, Johannes Ponge, Veronika Jaeger, André Karch.

Writing – original draft: Janik Suer, Veronika Jaeger, André Karch.

Writing – review & editing: all authors.

## Data and Code Availability

The code and data to reproduce the presented results are available at: https://github.com/JanikSuer/contact_structure_comparison. The agent-based model GEMS is available at: https://immidd.github.io/GEMS/.

## Appendix

### Contact Settings

Households are one of the most important contact sites for the spread of many infectious diseases, as household contacts are generally close contacts that often involving physical contacts, and occur over long periods of time [41,42]. Compared to other types of contacts, household contacts are difficult to avoid because the shared living space can rarely be completely separated. Therefore, especially during periods of severe contact restriction, such as lockdowns, household contacts are critical to the pandemic’s progression. This highlights the need for models to accurately reflect the household contact structures, especially for models that aim to investigate the effects of contact restrictions such as GEMS.

In GEMS households are determined by the synthetic population generation of Gesyland and are modelled without any substructure assuming a random mixing between all household members. Using the uncalibrated model parameter 𝜆_𝐻_ = 1 all individuals living in households with more than two members have on average one contact per day, while single households have none. This causes the average number of contacts in the household to be less than 𝜆_𝐻_. Since the contacts of individuals living alone are not affected by varying 𝜆_𝐻_ the average number of contacts still scales linearly with 𝜆_𝐻_ such that the calibration can still be applied.

Schools were subject to substantial restrictions in many countries. These restrictions were not enforced without reason as schools have been identified as significant contributors to the spread of pathogens [43]. Contacts in the school include both close-proximity indoor contacts and physical contacts ideal for the transmission of various pathogens. The age-structure of these school contacts in Germany is directly related to the class system, as most contacts occur in the school classes where only students of a similar age meet. However, during school breaks there may also be contacts between individuals of different classes and ages leading to a higher level of contact heterogeneity.

In GEMS the school is comprised of four substructures that include the school classes, school years, schools and school complexes. Here, the latter can always include multiple ones of the previous. This structure mimics the German school system, in which multiple schools frequently share a common schoolyard or other facilities, leading to interactions between students from different schools. Within schools, contacts between different schoolyears are facilitated, as the school years are often allocated to the same building. Contacts in the schoolyears arise independently of the class structure, e.g., through friendships that occur between different classes or lessons that include students from multiple classes. However, most contacts occur between individuals from the same class, as students spent a majority of their time in the same room as their classmates and form social groups in those classes. The structure of GEMS allows to capture all these contacts and specify their frequency individually. As in the COVIMOD data schools in GEMS also include kindergartens and universities.

Workplaces are locations where most adults spend a large proportion of their waking hours and have a large proportion of their daily contacts. Like children’s school contacts, workplace contacts often take setting over long periods of time, e.g., in shared office spaces and occur during different activities such as talking or eating together. The structure of workplaces is highly heterogeneous resulting in very heterogeneous contact behaviours. While some people are self-employed and work alone, others work in shared office spaces with shared cafeterias. The daily contact behaviours of these individuals differ significantly. Similarly, the contacts at work might be stable as in the case of office workers or vary from day to day as in the case of restaurant workers or clerks. Another influencing factor is remote working, which results in a complete physical separation of the people while still maintaining productivity. However, this is only not possible for all types of work.

In GEMS, workplaces, like schools, have four substructures. They consist of offices, departments, workplaces and workplace sites. Again, the latter includes multiple ones of the previous. The age structure in all substructures of the workplaces is similar, with no age-assortivity. While the substructures do not affect the age groups that come into contact with each other they do contain different pools of people that allow for different levels of mixing. Inidividuals who work in the same office will have more contacts with each other than individuals who work in the same workplace site (building) but have no other work relations. However, the pool of potential contacts is much larger for the workplace site leading to a more homogeneous mixing of the population. Therefore, the main aim of using the substructures for workplaces is to be able to represent different levels of mixing of the population. Since the investigation of the mixing levels beyond the scope of this work restrict the analysis is limited to the offices alone.

Other contacts in this analysis are all contacts that occur outside of the household, school and workplace. These contacts include a wide variety of interactions, such as random contacts in shops but also contacts with friends and family outside of the household. These types of contacts were the main targets of contact restrictions during the Covid-19 pandemic. However, it is almost impossible to completely ban these other contacts, as they include contacts that arise during essential activities, such as going to the supermarket or to the doctor.

While the COVIMOD dataset includes further distinctions of the contact type, such as shopping activities or sports, we have aggregated these types into other contacts to allow the comparison with the contacts generated in GEMS. GEMS uses an individual’s municipality as the location for other contacts. This can represent random contacts between individuals in supermarkets or waiting rooms but cannot capture reoccurring contacts such as regular meetings with friends. This approximation is used to maintain the static association of individuals with settings while also keeping the number of settings small enough to be efficiently handled.

### Substructure Contact Matrices

**Figure 6:**
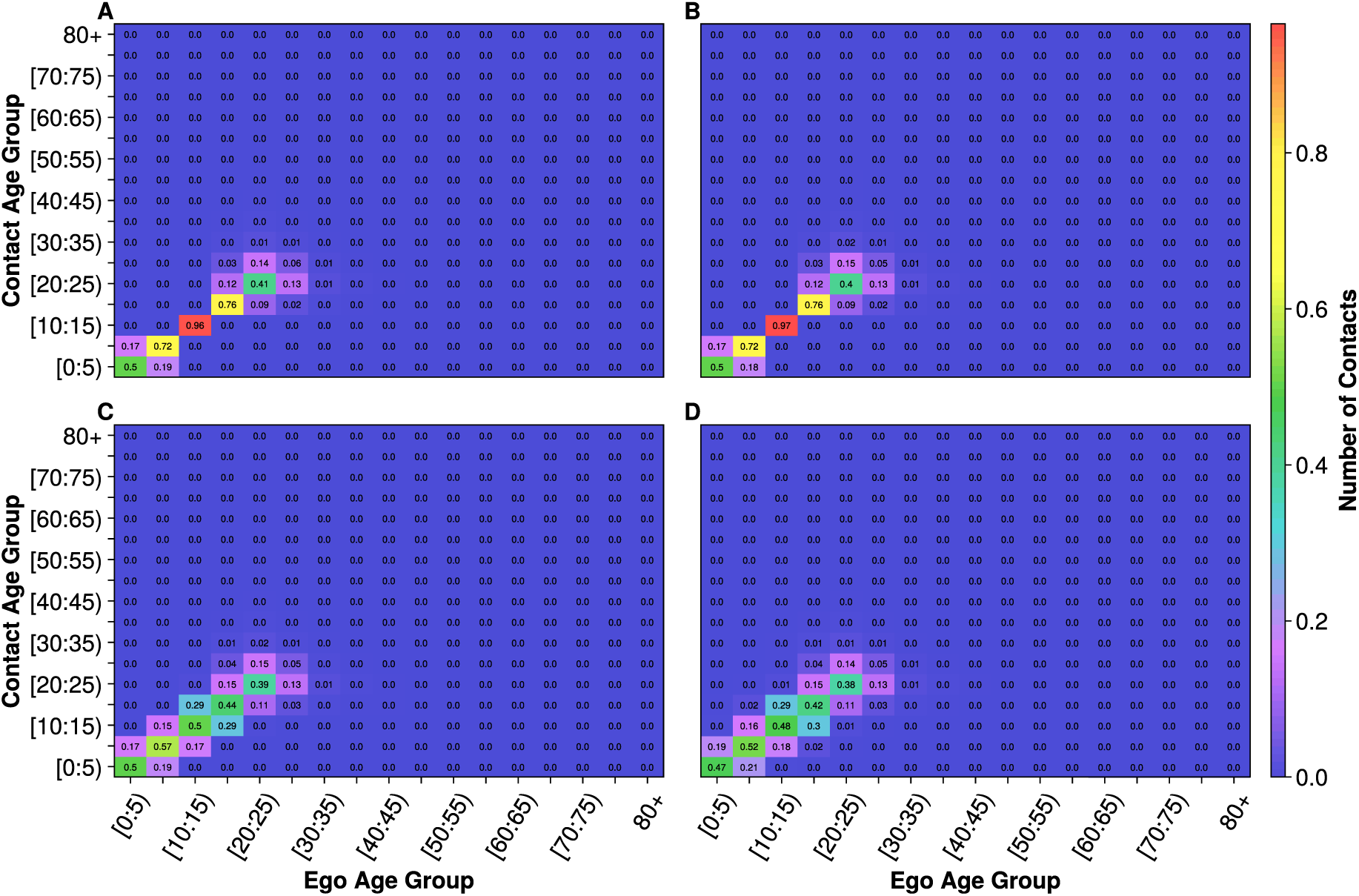
Age x age contact matrices for all hierarchical levels of the school including the (A) school class, (B) school year, (C) school and (D) school complex in GEMS. Note that the school classes and years as well as the school complex and school exhibit have similar age structure, respectively such that the contact structures are almost identical.

**Figure 7:**
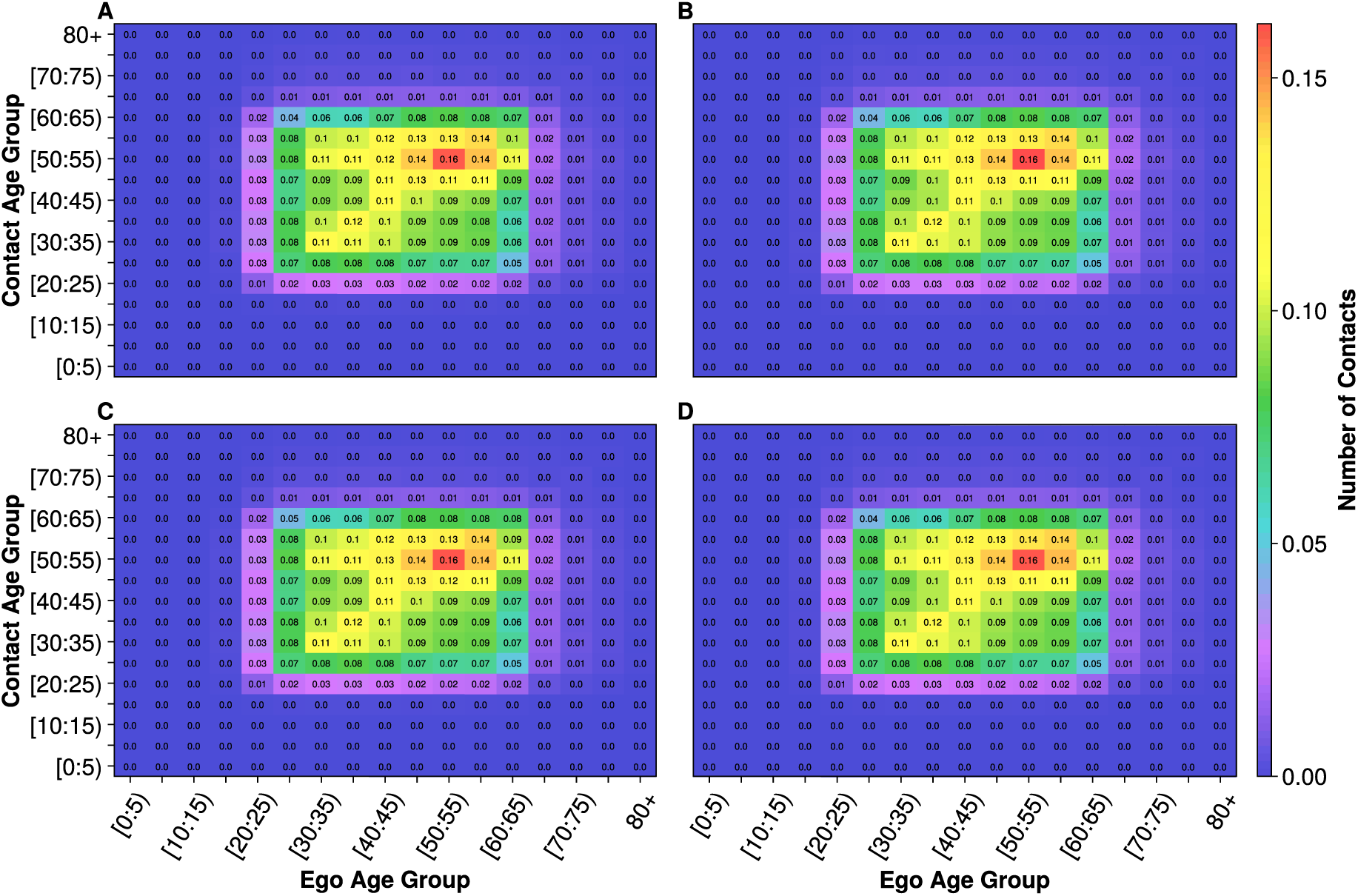
Age x age contact matrices for all hierarchical levels of the workplace including the (A) office, (B) department, (C) workplace and (D) workplacesite in GEMS. Note that the age structures of all hierarchical levels of the workplaces are similar, such that the contact structures are almost identical.

